# “We’re here to help them if they want to come”: A qualitative exploration of hospital staff perceptions and experiences with outpatient non-attendance

**DOI:** 10.1101/2024.09.14.24313685

**Authors:** Shayma Mohammed Selim, Steven M McPhail, Hannah E Carter, Christina Malatzky, Sanjeewa Kularatna, Sundresan Naicker

## Abstract

**Background:** Patient non-attendance remains a major challenge for health services. Few studies have examined how health service providers think about, potentially address, and prioritise non-attendance within the scope of their practice. This study aimed to (1) explore healthcare professionals’ perspectives, beliefs, and opinions about the impact of patient non-attendance within a publicly-funded outpatient physiotherapy clinic context; (2) explore perceived barriers and facilitators associated with the implementation of non-attendance mitigation strategies; and (3) identify health service staff generated solutions to address perceived barriers and enhance facilitators.

**Methods:** A focus group discussion and semi-structured interviews were conducted between June 2023 to January 2024 with 27 physiotherapy department clinic outpatient staff involved in operationalising clinic referral processing, appointment scheduling, or providing care to patients. Data was analysed using a hybrid inductive/deductive framework analysis approach.

**Results:** Participants indicated that non-attendance had predominantly negative implications for the health service, healthcare provider, and patient. The interconnected issue of non-attendance encompassed multiple areas and were broadly categorised into five inductively identified themes: impact of non-attendance, perceptions of value, material deprivation, service delivery and built environment, and professional role and identity. Non-attendance mitigation strategies generated by participants were deductively mapped to the theoretical domains framework (TDF) to explore behavioural determinants that may influence successful implementation. This included knowledge, reinforcement, goals, optimism, memory, attention and decision-making, environmental resources and context, and emotions.

**Conclusions:** Staff identified multiple strategies for reducing non-attendance; implementing many of these strategies would require additional resourcing. Research determining the effectiveness of such strategies both in the short-term and long-term following implementation into practice remains a priority for future investigation.

## Introduction

Unplanned non-attendance at outpatient appointments negatively impacts patients by contributing to delayed or missed treatment and negatively impacts healthcare services through missed reimbursement opportunities.^1,2^ Non-attendance can also contribute to ineffective and inefficient use of clinician time due to an inability to redirect time towards other productive tasks.^3^ While addressing non-attendance remains a significant challenge within modern healthcare systems, there is a general belief that a one-size-fits-all approach for mitigating non-attendance does not exist.^4^ Rather, mitigation strategies should be tailored to the scale and specific needs of healthcare systems, providers, and the patients they serve.^4,5^

Several factors are known to be associated with non-attendance, often relating to patient-level and system-level determinants, such as patient forgetfulness, scheduling conflicts, miscommunication between the service and the patient, and many other complicated scenarios.^6^ Research has often focused on evaluating reasons for non-attendance from the patients’ perspective.^7,8^ However, there is a paucity of research examining how health service providers think about, potentially address and prioritise non-attendance within the scope of their practice.^7–9^ Some evidence suggests that perceptions and related evaluations about the reasons for non-attendance influence how health service providers may use and consider mitigation strategies.^10^ In the absence of such information, qualitative exploration may facilitate the analysis of nuanced perspectives.^11^

Physiotherapy plays an increasingly important role in facilitating the prevention and management of acute and chronic health conditions.^12,13^ As a consequence of population growth and ageing, demand for physiotherapy services is expected to continue to grow and can span the provision of a range of specialised treatments in areas such as geriatrics, sports physiotherapy, women’s health, and musculoskeletal physiotherapy, among others.^14^ In contexts such as Australia, publicly-funded physiotherapy services have often been deemed essential and effective for almost 7 million people with musculoskeletal conditions (e.g., osteoarthritis or back pain).^14,15^ In 2020–21, healthcare expenditure for these conditions was estimated to be AU$14.7 billion, representing the country’s highest spending of all disease groups.^15^ Globally, musculoskeletal disorders are estimated to affect more than 1.6 billion people.^16^ As such, patient non-attendance remains an ongoing concern within this clinical context because evidence suggests that the targeted specialist outpatient waiting list timeframes are often exceeded, which could potentially be detrimental to patient health. It may further contribute to decreased patient satisfaction and lead to sub-optimal healthcare service delivery.^17^

This study had three principal aims: (1) explore healthcare professionals’ perspectives, beliefs, and opinions about the impact of patient non-attendance within a publicly-funded outpatient physiotherapy clinic context; (2) explore perceived barriers and facilitators associated with the implementation of non-attendance mitigation strategies; and (3) identify health service staff generated solutions to address perceived barriers and enhance facilitators.

## Methods

### Study design

This cross-sectional qualitative study was conducted with a focus group discussion and semi-structured interviews. The Consolidated Criteria for Reporting Qualitative Research Checklist (COREQ) was used to guide the transparent and complete reporting of study methods (See Supplementary Material).^18^ This study received ethical approval from the Metro South Health Human Research Ethics Committee (HREC/2021/QMS/81605) and written informed consent was obtained from all participants.

### Setting

This study was conducted at three physiotherapy outpatient clinics at two large tertiary metropolitan hospitals in the Metro South Hospital and Health Service (MSHHS) region in Queensland, Australia. The MSHHS serves an estimated 1.2 million people in its designated catchment area.^19^ The first participating clinic (Clinic 1) specialises in vestibular, rehabilitation, and musculoskeletal physiotherapy. The second clinic (Clinic 2) specialises in women’s, men’s, and pelvic health physiotherapy, and the third clinic (Clinic 3) specialises in only musculoskeletal physiotherapy. Each of these clinics are publicly funded and free to access for Australian residents with eligible referrals.

### Methodological approach

This study used an implementation science methodology to inform the overall approach, which was organised according to the following steps: initial context assessment including a stakeholder discussion and document review; triangulation of findings to inform an initial set of semi-structured interview questions; conducting one-on-one interviews and a focus group discussion; iteration and refinement of questions to ensure richness of data^20^; inductive analysis of the interview and focus group data using an interpretive description (ID) approach, and deductive framework mapping to the theoretical domains framework (TDF) adapted from the Atkins et al. methodology.^20–23^ The approach followed is presented in Figure 1.

**Figure 1.**
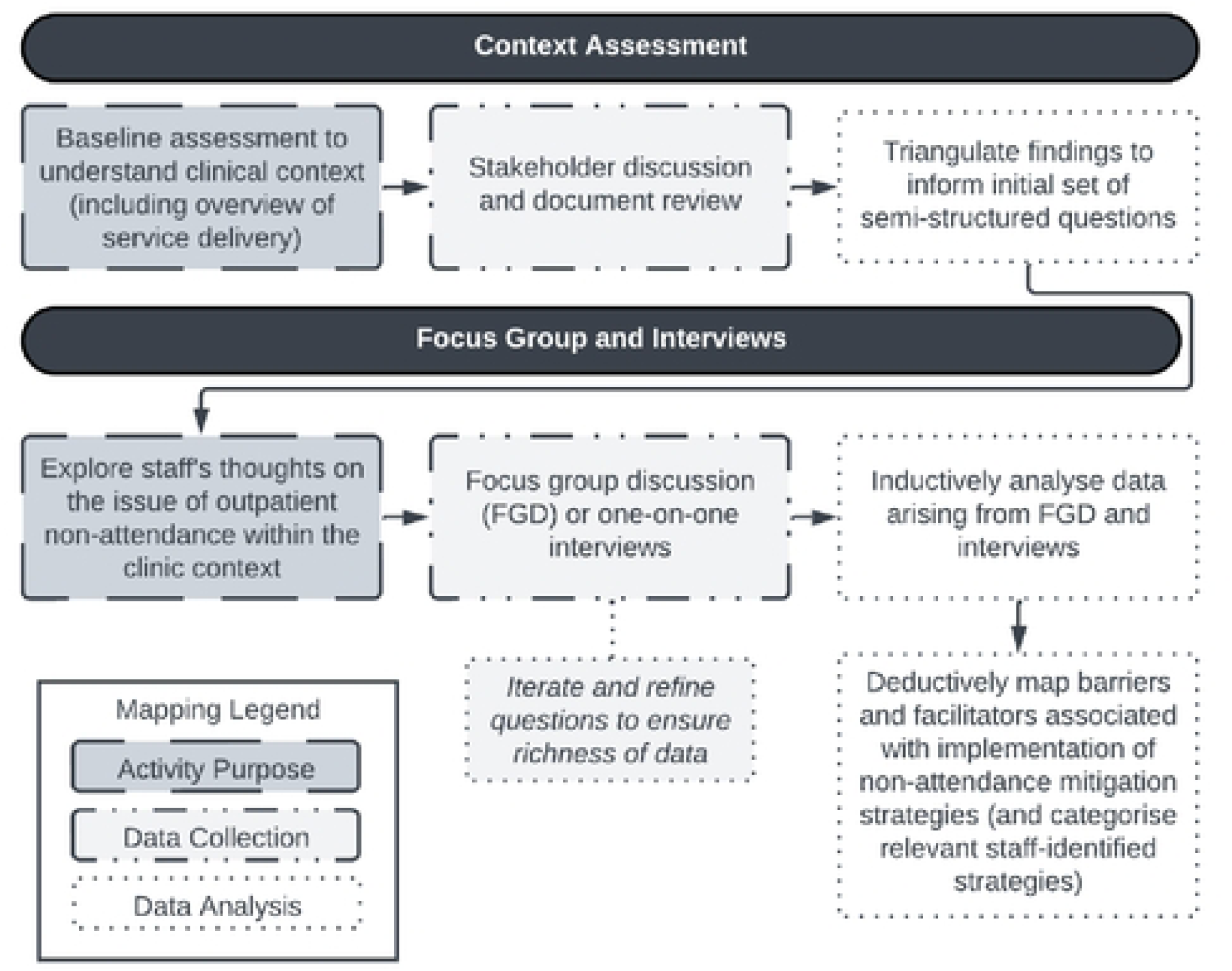
Flowchart of methodological approach

### Inductive approach using interpretive description (ID)

To understand healthcare professionals’ lived experience of patient non-attendance, an interpretive description (ID) approach was used.^22^ This approach primarily focuses on capturing and interpreting participants’ views surrounding the meaning of a particular experience under inquiry (i.e., a phenomenon).^22^ Additionally, using an ID approach, qualitative inquiry aims to develop contextually-relevant knowledge that may inform improvements in healthcare service delivery.^22^ Physiotherapists and administrative staff both encounter non-attendance, often on a day-to-day basis. Hence, understanding their perspectives of this phenomenon is crucial to providing insights into prospective improvements to non-attendance mitigation strategies. This approach thus informed the inductive analysis of data, which then formed the basis for deductively mapping participant-identified approaches for mitigating non-attendance to the theoretical domains framework (TDF), a well-established theoretical framework of behaviour change.^23–25^

### Deductive approach using a behaviour change framework

Implementing acceptable and suitable interventions within healthcare requires changes in behaviour on an individual (i.e., those who use the intervention) and collective (i.e., the system in which the intervention is used) level.^23^ Facilitating behaviour change is a complex activity that requires understanding the influences on behaviour in the context in which it occurs.^23,24^ The TDF provides a structured, theory-based approach to understanding healthcare professionals’ perspectives of the contextually relevant barriers and facilitators to change associated with implementing non-attendance mitigation strategies and a mechanism to categorise staff-generated solutions against behaviours to target that are integral to successful implementation.^23,26^ Deductive themes, as identified using the ID approach, were mapped to TDF domains where relevant to identify described mitigation strategies and related barriers and facilitators.^27^ This two-tiered inductive-deductive approach allowed for analytical flexibility, as recommended by McGowan et al.^27^.

### Study participants and recruitment

Physiotherapy department clinic outpatient staff involved in operationalising clinic referral processing, appointment scheduling, or the provision of care to patients were purposively sampled to ensure a mix of junior and senior staff perspectives were represented. This included administrative staff, clinic managers, and physiotherapists. Potential participants were invited by a member of the research team, who is not in an unequal relationship with potential participants (but may be known to some participants) to participate in a context assessment, one-on-one interviews, and/or focus group discussion via email. Participant information sheets and consent forms were emailed with the invitation to participate. Based on ID methodological recommendations, the final sample size was not determined by data saturation.^22^ Instead, an estimated sample size of 25 to 30 participants was initially established to enable analytical depth.

### Data collection

#### Developing interview materials

An initial discussion with key stakeholders was conducted to clearly define and explore the issue of outpatient non-attendance within the physiotherapy outpatient context. These key stakeholders included a clinic director (n=1), physiotherapist clinic managers (n=2), and an administrative manager (n=1). The discussion centred on understanding relevant work processes and latent protocols as they related to the experience of non-attendance within their practice so that the interview guide questions were contextually relevant to participating physiotherapy clinic staff. Topics were developed to explore participants’ thoughts on the impacts of non-attendance, perceptions of the barriers and facilitators to patient attendance, and opinions about current or prospective non-attendance mitigation strategies (See Supplementary Material).

#### Research participation and facilitation of focus group/interviews

Data collection via focus group discussion and one-on-one semi-structured interviews was conducted between 01 June 2023 and 31 January 2024. Based on participants’ preferences and availability, participants were provided with the option to participate in either a one-on-one interview or a focus group with up to 8 other participants. Both could be conducted face-to-face or virtually (videoconference via Microsoft Teams), with the exception of the additional option of virtual delivery via telephone for interviews. Interviews were approximately 45-minutes in length, while the focus group was conducted for approximately one-hour. The interviews and focus groups were audio or video recorded (depending on the interview media used), with informed consent obtained prior to commencement and transcribed verbatim. The focus group discussion was led by SMM, an experienced health services researcher and clinician (male, PhD qualified, embedded in the health service and known to some participants). SN, an experienced mixed-methods health services researcher and implementation scientist (male, PhD qualified), and SMS, a female PhD student with a research focus on health economics, were also present during the focus group. All semi-structured interviews were conducted by SMS. Participation was voluntary, and participants could withdraw without penalty at any time during the data collection process. Consistent with an interpretive description approach, member checking, a form of post-interview validation in which participants have the opportunity to review, comment, or change their responses, was not used. This may potentially limit the risk of interpretations being swayed and for the formation of meaningful clinical implications being impeded.^22,28–31^ As described below, data from the interviews and focus group discussion were reviewed and analysed iteratively to allow recruitment to cease when the research team felt an adequate and deep understanding of participants’ experiences with the phenomenon under inquiry was achieved.^22,32^

### Data analysis

To address the study aims, we applied a hybrid inductive/deductive framework analysis approach for data analysis.^21,33^ Data analysis was conducted using NVivo software (release 1.6.1).^34^

### Inductive analysis

Firstly, we used an inductive analysis approach that was concordant with an ID methodology. The focus group and interviews were transcribed verbatim and de-identified. Transcripts were checked for accuracy by SMS and were corrected where necessary. To become immersed in and familiar with the data, members of the research team (SMS and SN) thoroughly read through each transcript and listened to the corresponding audio/video recordings. One interview transcript was then broken down into portions by SMS and SN, with each portion representing a distinct opinion or concept. Representative quotes were extracted for each of these portions where appropriate. Information was then organised into distinct sub-themes, and these were classified into higher-order themes, or general concepts. This process was repeated for each transcript, and a series of discussions were held between the two researchers to discuss and further develop the arising concepts and subthemes identified. Throughout this process, caution was taken to ensure that derived concepts authentically reflected the participant’s voices.

### Deductive analysis

As described above, barriers and facilitators associated with the use of non-attendance mitigation strategies were inductively identified from the data. These were then deductively mapped into one of the 14 theoretical domains of the TDF to inform categorisation of staffs’ suggestions for future intervention options.^24^

### Rigor

The research team sought to be open and reflective throughout the data collection and analysis stages of this study about the influence of the researchers’ subjectivities and preconceived assumptions on the research process.^22,35^ This included critically assessing what the research team already knew or expected to find out about how patient non-attendance impacts and is experienced by healthcare professionals. This process was facilitated using a reflection journal in which thoughts, questions, or ideas arising during data collection and analysis were recorded and were then brought into team discussions. During the focus group and interviews, the facilitator consciously made the effort to provide space for participants to explain and elaborate on their views. This aimed to ensure that a diverse range of views were captured. This process involved using open-ended questions, avoiding leading questions, and adopting a non-judgmental approach to interviewing.^36,37^ Additionally, the facilitators conducted regular debriefing sessions during data collection to clarify thoughts, and where necessary, adjust interview questions.

## Results

### Participant characteristics

A total of 27 consenting participants took part in the study: focus group (n=1 (with 8 participants, 30%)) and interviews (n=19, 70%). Most participants were physiotherapists (n = 22, 81%) and belonged to one of three clinical divisions as described in Table 1. All administrative staff worked within the allied health service department and had experience working across multiple disciplines including but not limited to physiotherapy, occupational therapy, and dietetics.

**Table 1.** Participant characteristics.

| Characteristics | Number of participants<br>(n = 27) |
| --- | --- |
| **Sex** |  |
| Male | 9 (33%) |
| Female | 18 (67%) |
| **Role** |  |
| Admin | 5 (19%) |
| Physiotherapist | 22 (81%) |
| **Physiotherapy Division\*** |  |
| Vestibular, rehabilitation, and musculoskeletal | 8 (30%) |
| Women's, men's, and pelvic health | 7 (26%) |
| Musculoskeletal | 7 (26%) |
| *\*division only applicable to physiotherapists* |  |

### Themes

Five major themes were inductively identified from the data as described in Figure 2.

**Figure 2.**
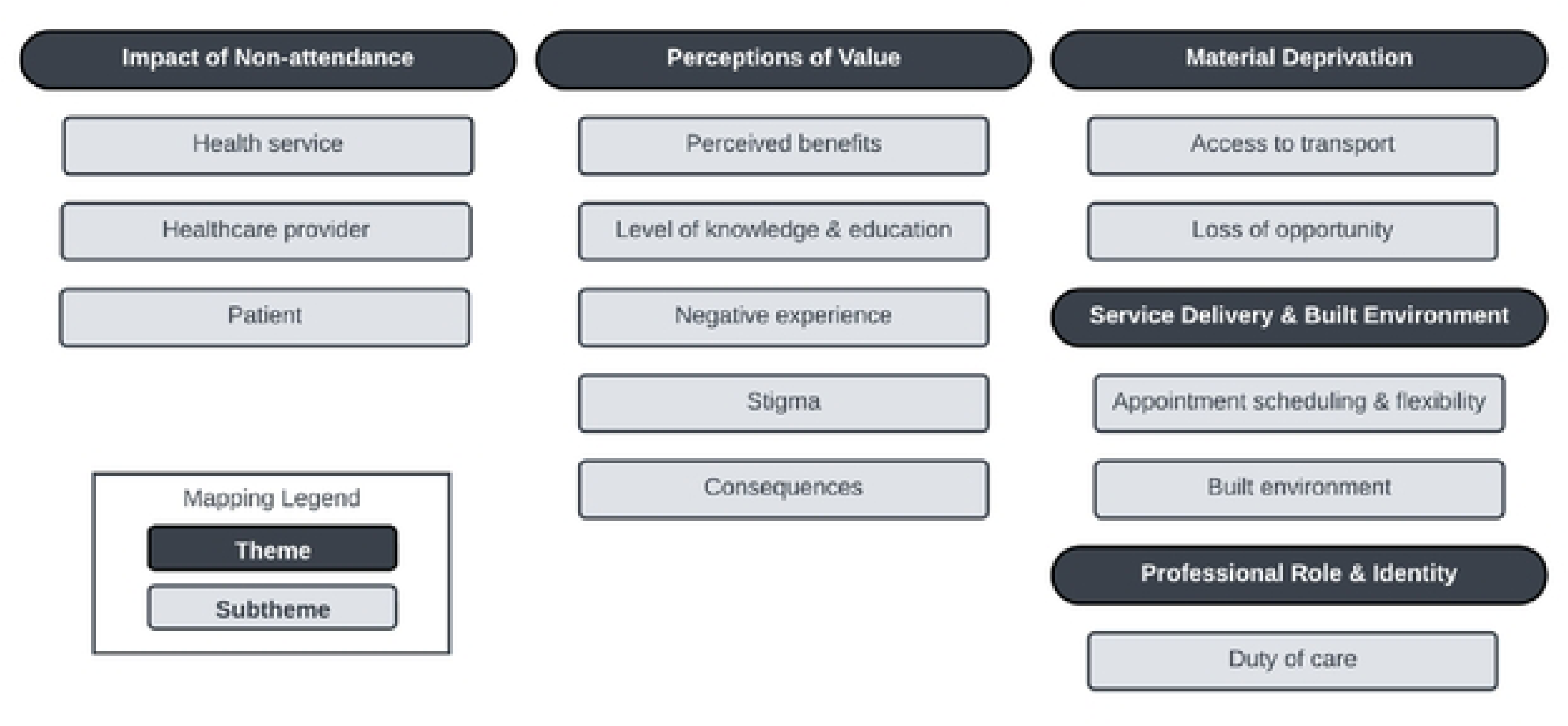
Thematic schema of physiotherapy staff perspectives surrounding patient non-attendance

### Impact of non-attendance

This theme explains the extent to which non-attendance affects the health service, healthcare provider and the patient as described by staff.

### Impact on the health service

The inability to get a patient into a vacant appointment slot was deemed the key concern associated with non-attendance at the health service level. This had implications for other patients on the wait list who may have lost the opportunity to be seen at an earlier date.

> “We’ve got wait lists and we can’t get those patients in… So yes, it definitely… impacts the service, but it also impacts all of the patients that are waiting that want the appointments.” – P11 (Admin)

In addition, non-attendance may lead to loss of revenue to the department, and the potential inefficient use of resources resulting from patients requiring further management in the future to address the recurrence of symptoms.

> “I think it’s lost income, it’s lost opportunity for the patient and for the clinician.” – P15 (Physiotherapist)

> “…at the end of the day, a complication or a recurrence will cost more to the service.” – P12 (Physiotherapist)

### Impact on healthcare providers

For healthcare providers, including both administrative and clinical staff, there was consensus that non-attendance often had differential impacts on workflow. In some instances, patient non-attendance enabled clinical staff to catch up on other work or rest during a busy shift.

> “And then sometimes also, like, a selfish personal thing is like, if you’re overbooked or like, feel really stressed and you have other things, sometimes it’s a breath of fresh air. I feel like ‘oh my god, great, I can catch up with all my other notes’” – P14 (Physiotherapist)

In other instances, clinical staff described how non-attendance significantly increased the workload and decreased the efficiency of service delivery. When patients miss appointments, it could disrupt the continuity of care, making it difficult to consistently schedule them with the same clinician. As a result, new clinicians may need to spend time relearning a patient’s history and treatment plan, which may often require re-evaluation based on their experience. This not only consumes valuable staff time but can also lead to the provision of treatment plans that may not align with patient preferences, ultimately impacting the overall quality of care.

> “One of the things that I find that impacts on us is… when we have a patient… we try to see the same patient, we understand what is going on and we learn this patient, how they usually like to be treated, how they respond to therapy, and how we can progress them. When we have to change their appointments and they… reschedule with another clinician, the new clinician has to have all that learning process again, which… takes time… The other thing… is that the patient, because of any changes of clinicians or time wasted, is also potentially going to take a little bit longer to recover. So, it has an impact on the patient as well because if you see the same clinician, we really know what is going on.” – P12 (Physiotherapist)

Similarly, non-attendance impacted administrative staff by increasing the number of tasks required to facilitate patient attendance. One participant described this experience as,

> “It does create work for us. If the patient was to show up, we would simply make them another appointment if they needed one or discharge them, whereas if they fail to attend, we have to cancel their appointment, put an appointment back on the waitlist, send the patient a letter. Then in two weeks’ time, check to see if they’ve made an appointment, and if they haven’t, send them another letter to let them know that they’ve been discharged.” – P11 (Admin).

### Impact on patients

Staff believed that non-attendance impacted both new and current patients. For patients who were still on wait lists, non-attendance potentially contributed to delayed access to care. In contrast, for patients who were already seeing the service and failed-to-attend, continuity of care was often hampered. Vacant appointment slots that aren’t able to be filled could have potentially been allocated to patients who may benefit from earlier access to treatment. This delay in access to care may lead to their condition not improving or plateauing, which could have ramifications for a patient being able to return to work or how they function day-to-day.

> “…puts risk on the patients because patients can’t get treated in a timely manner anymore because the blank spots are taken up by no shows.” – P03 (Admin)

In instances where there are big breaks in care resulting from inconsistent appointment attendance, reinforcing appropriate adherence to treatment was often considered challenging because there may be a higher likelihood of patients forgetting how to do the prescribed treatment plans (e.g., exercise).

> “…it just prolongs the… rehab times by not being able to progress exercise plans and treatment as we would like and as based on evidence-based practise would suggest.” – P16 (Physiotherapist)

### Perceptions of value

This theme explores how healthcare professionals think about patients’ views regarding the value (i.e., the usefulness or importance) of attending scheduled clinic appointments.

### Perceived benefits

There was consensus amongst staff that a contributing factor to attendance was whether patients perceived positive benefits from attending. These benefits were varied and could include seeing improvements in their condition, the ability to ward off social isolation through interaction with clinicians, and feeling a sense of ownership in addressing their healthcare concerns. One participant described their thoughts on the perception of benefit as,

> “I think for patients that continue to come it’s usually because they are seeing a benefit from doing it, which is kind of the way we work in society, if we see a benefit, we keep doing something, if we don’t see a benefit, maybe we won’t do that so much anymore.” – P02 (Physiotherapist).

Staff indicated that therapeutic rapport between the patient and provider was a key component affecting patient engagement with the health service. One participant summarised this as,

> “they’re feeling positive and good about their healthcare experience thus far, so that sort of helps set a positive mindset towards continuing health care.” – P10 (Physiotherapist).

For some participants, there was belief that patients who often felt socially isolated significantly valued the interaction they would receive by attending an appointment, particularly because it gave them a platform from which to be heard.

> “There are… some that feel like there’s someone to talk to… if they don’t have close family, friends, they live alone, they see the physio, even if it’s only once a month… they get to come out of the house… they come and see someone… they have a chat to someone.” – P02 (Physiotherapist)

However, staff also highlighted that though it could feel rewarding to see that patients are engaging with the health service, this could become difficult to navigate when patients are no longer clinically progressing but do not want to be discharged from the service because they would lose the ability to continue to see someone who listens to them.

> “…despite finishing treatment… not seeing any further effect… they decline to be discharged… I usually give them opportunities. I won’t see them as quick as I probably will with another patient, but I will book them for a month or six weeks and say, ‘OK, this is going to be your last review’, and it comes to the last review and they’re still not (ready).” – P12 (Physiotherapist)

Patient readiness, characterised as the state in which a patient is ready to receive and engage with a care plan at the point at which they are given an appointment, was deemed as an important consideration relating to the prospect that a patient may place value in attending an appointment. Whilst patients may indicate to administrative staff that they are available to attend a scheduled appointment during the booking process, they may not necessarily be ready to receive care at that time for several different reasons. This may include but is not limited to being mentally prepared to accept and accommodate the need for following treatment plans for their condition.

> “…some people aren’t ready to engage or its they can’t quite engage because they probably need more mental health support… financial support, transport support.” – P05 (Physiotherapist)

Some staff highlighted that accountability and ownership of one’s health condition/s and initiative to engage with care needs are up to the patient as autonomous agents and that healthcare providers aim to cater to patient needs when a patient is ready for care.

> “It’s also, like, ready to initiate… almost somebody that’s like, ‘I’m going to have to do these things’… ready to like, get to the headspace of, ‘I accept that I need to also do all of these things… for me to see treatment outcomes’.” – P14 (Physiotherapist)

> “It has to be patient-driven, and we’re here, ready to help them if they want to come.” – P06 (Physiotherapist)

Some staff indicated the difficulty of instilling motivation in patients to attend when there is a lack of patient ownership and the trickiness of determining how best to mitigate non-attendance in such cases.

> “You could try to motivate someone and educate someone to like the cows come home, but if they don’t wanna like, find that within themselves, you can’t like force out upon somebody. They have to be ready to do physio, to want to participate.” – P04 (Physiotherapist)

> “I think the ownership should be… put back on the patient. Our job shouldn’t be going around chasing… after them – why aren’t they turning up to the appointment. And they’re getting a lot of reminders already, like they get a text message, they get like a phone call, and then normally we send the letters. I’m not sure what else we can do.” – P06 (Physiotherapist)

### Level of knowledge/education

Staff indicated that the perception of treatment benefits was linked to patients’ level of knowledge about potential positive health outcomes relayed to them by their referrer and that without clear communication from referrers, there may not be a strong enough incentive for patients to attend appointments.

> “Often the seed is planted in primary care by the GP… in their mind they think they need to have some sort of surgical intervention… we provide education from day one… to challenge those beliefs, and for some people it’s a relief, they don’t want surgery. So, they’re like, ‘oh, great, you mean I don’t need surgery’. So, they’re very motivated.” – P09 (Physiotherapist)

Some staff highlighted the importance of patient attendance at an initial appointment, which provides the space to educate the patient on the benefits that physiotherapy could offer them. However, this becomes difficult because of the uncertainty surrounding what the patient has been told by their referrer. One participant described this experience as,

> “… it depends on who’s the primary contact practitioner at the time. I think that is somehow out of our control… when they’re here, we can educate them for sure, but you gotta go back to the source…. It’s up to the medical doctor or practitioner to make sure they educate them.” – P06 (Physiotherapist).

### Negative experience

Negative previous and ongoing experiences with healthcare providers, such as poor communication, can deter patients from attending appointments. This includes experiences in primary care and physiotherapy care and can range from patients not receiving enough information throughout their health journey such as who they are being referred to and why they are being referred to that service; having experienced poor quality clinical care; and a lack of proper engagement with patients to ensure that they are being provided with sufficient information to decide on care that they feel is appropriate for them.

> “… they may already have tried physio and I know there’s variation between… how different physios manage the same condition – they might have a bad experience from physio. Physio hurt them so that’s why they don’t wanna come.” – P06 (Physiotherapist)

Staff indicated the importance of building rapport with patients to combat non-attendance attributed to prior negative experiences. One participant summarised this as,

> “…I think rapport… like if they (patients) have trust and have built a rapport with their clinician that will help with attendance because then I suppose you get the ‘I want to go to my appointment to get whatever advice, education, exercise, etcetera from it’. But in a way, and this is like personally too, like, I don’t want to miss some of my appointments because I’m like, ‘I don’t want to let the person down’”. – P14 (Physiotherapist)

### Fear of stigmatisation

Staff indicated that patients may avoid seeking care because they have had prior or ongoing negative experiences, such as being stigmatised by healthcare providers, which may have resulted in the patient becoming fearful of engaging with a health service.

> “… lots of patients come in with a lot of psychosocial stuff… patients who come in and they’re in chronic pain for years and there’s no identifiable injury… they’ve got pre-contemplation and their fear avoidant… they’re fearful of a lot, of doing a lot of activity, fearful of engaging in activity, fearful of exercise” – P09 (Physiotherapist)

Other staff expressed that some patients may feel embarrassed to attend if they haven’t completed the required treatment plan. One participant summarised this as,

> “…they’re either disappointed in themselves or too scared to like tell us, or they just haven’t done their exercises or what we’ve asked so they just don’t come” – P04 – Physiotherapist.

### Consequences

Some staff expressed that the lack of consequences for failing to attend appointments may play a role in the reduced accountability held by patients. This includes perceptions that the ability to easily reschedule appointments following non-attendance contributes to patients not placing value in attending appointments at the scheduled time.

> “I think it still kind of does come down to, we learn from consequences or we learn from action and so if we keep forgetting but we can still get the opportunity to come, then you’re not ever really learning from those behaviours… I think that that also is kind of giving an indication of how much value they’re really putting on the care that they’re getting too.” – P05 (Physiotherapist)

Additionally, some staff contemplated the introduction of a monetary fine for non-attendance but acknowledged the use of such consequences would not be feasible within publicly-funded healthcare systems where services are often covered by Medicare, and which are aimed at providing equitable access to healthcare based on need as opposed to ability to pay for services. However, staff acknowledged that this practice may often be adopted for privately-provided services.

> “I wonder if that sort of forgetfulness happens a bit more frequently because… if they don’t come, we give them a call, we might give them… a second chance… to come back and it doesn’t seem to the patient like there’s been any… personal consequence versus maybe like a private appointment where it’s a fee paying service and failing to attend incurs a 50% fee or a $20 fee or something… patients would, I don’t know, feel that personal impact a bit more.” – P10 (Physiotherapist)

> “I think if people had to pay, they probably would be more inclined to come, but I don’t necessarily think that’s ever going to be feasible nor do I necessarily feel like that’s ethical in the public health system…” – P10 (Physiotherapist)

### Material deprivation

This theme describes how staff think about and perceive the impact of limited resources and related financial constraints on patients’ ability to attend scheduled appointments.

### Access to transport

Staff indicated that the inability to access transportation is one of the most direct means by which material deprivation can impact a patient’s capacity to travel to healthcare providers for scheduled healthcare appointments. This can be particularly challenging for patients who do not live within the vicinity of the clinic or in areas with limited public transportation options.

> “For our demographic… some of them say they can’t physically afford to get here… there’s no money on their go card. They can’t afford… public, private transport.” – P11 (Admin)

> “Some people don’t have anyone who they can ask to come and drop them off” – P12 (Physiotherapist)

For those patients who may be able to drive to the healthcare provider for the appointment, the lack of free parking may be a potential deterrent for some patients.

> “…there’s no free on-site car parking, they have to pay… being able to come here is like, ‘oh, either I come to my hospital appointment and have to pay for parking, but I don’t eat… I don’t have a day’s worth of food or something like that’. Like people are going to pick the cost of living over… having to pay to come to an appointment.” – P02 (Physiotherapist)

Patients who may experience material deprivation may also have limited access to information about support programs. This could be attributed to a lack of awareness by referrers or treating physiotherapists about schemes that their patients may be eligible to access or which patients may need to access such services.

> “…the patients, they don’t have any education on the community transport that’s available – so patients can go to their GPs and they can organise either very low cost or free community transport to bring them into the hospital. But most of them don’t know about that… or they don’t wanna bother anybody. So, then they just don’t come for their appointments because they can’t get here.” – P11 (Admin)

### Loss of opportunity

Patients experiencing material deprivation may have competing responsibilities or priorities such as work or family commitments. This can often make it challenging to allocate time towards attending scheduled appointments.

> “…which is again, what I played to in terms of where they rate physio in terms of importance compared to everything else that’s going on in their life. If they think it’s more important for them to earn money and work rather than attend physio.” – P16 (Physiotherapist)

> “Like, you know, got 5-6 kids and got no childcare… they don’t have family support… and they can’t leave with anybody they trust, I guess.” – P06 (Physiotherapist)

> “This is also probably important also, patient… they had a major trauma and they’ve already write up their sick leave – the fear of losing their job – probably under pressure to go back to work, that’s really important and they can’t avoid, especially if the appointment time doesn’t work for them work wise.” – P06 (Physiotherapist)

### Service delivery & built environment

This theme explores the impact of service delivery and the structural layout of healthcare services on patient attendance.

### Appointment scheduling and flexibility (appointment times and types)

Staff recognised that scheduling appointments during work hours (e.g., 9AM—5 PM) was a key limitation that may inhibit a patient’s ability to attend appointments. Some staff further indicated that they have often received requests from patients asking about the availability of weekend or after-hours appointments.

> “…we try and accommodate… as much as possible, and obviously does get to a point of, like, ‘well, if you really need out of hours type timeframes then the public service is probably not for you. You probably do need to go privately and see someone that way.’ But, if you can kind of manage it that, like, you know, as much as we can within the constraints we have within our service, we can see you and maybe every second or third appointment, like, we were just saying before, it is the face-to-face and in-between time, we do a phone call, if that’s going to mean your compliance and your ability to be able to attend appointments and make that more manageable. That does seem to make it worthwhile.” – P02 (Physiotherapist)

> “We open Monday to Friday, we open at 7:00 and we close at 4:30… I think if they were going to open later, it would need to be till like 7:00 at night so that patients could come after work or Saturday morning appointments… that would solve a lot of the problems. – P11 (Admin)

There was agreement amongst staff that offering alternative appointment types, including telehealth instead of face-to-face appointments, may be a good option to enable attendance, particularly for patients who may find it difficult to balance other work, healthcare or family commitments. However, within the physiotherapy discipline, providing remote appointment options may not be suitable as hands-on assessment and treatment may be required.

> “…phone calls or telehealth are always a good option, not necessarily for every patient or for every appointment. But… there’s lots of appointments that we can do it for, the phone calls can sometimes be really appreciated, especially… if they have transport issues or live far away.” – P03 (Admin)

> “I think sometimes trying to book their appointment on the same day that they’re seeing the doctor, some like you know, they’re going to be attending so they’re going to come to both are all little things that I’ll try to do to just make it more convenient for patients to come into the hospital.” – P04 (Physiotherapist)

### Built environment (mobility and navigation)

The built environment and structure of healthcare facilities specifically, can greatly influence patients’ ability to attend appointments. Patients with mobility issues or other disabilities may find it difficult to navigate through environments with inaccessible entrances and relatively long distances to their destination.

> “Our patients are physio patients – it is a hike from the car park to us. So, I had a patient who had [the pain management course], he got here, realised there was no parks anywhere near a door and rang us and said ‘I’m not coming because I can’t physically walk the distance from the car park to the clinic’. So he left.” – P11 (Admin)

Additionally, the structural layout and displayed signage within healthcare facilities may impact a patient’s ability to find their appointment locations.

> “It’s a real rabbit hole of trying to find your way around and how to get from A to B, and honestly, there are certain parts of the hospital, I have no idea how you get to… So, I can hardly imagine as a patient coming in… last time they were here, it looked like this and they knew how to get somewhere and then all of a sudden next time they’re there, it’s completely different… I imagine, for some people… [not] being able to find where you need to be and when you need to be there by, that could possibly be a little bit off putting and might be a bit daunting for some patients as well in terms of access.” – P02 (Physiotherapist)

Improving information on how to get to an appointment location or introducing digital maps that are accessible, interactive, and easy to understand may help patients find their way around the facility. This could reduce the likelihood of patients getting lost or arriving late for appointments.

> “Some people don’t know their way to get into hospital and they don’t know that probably there are quicker and shorter ways to get into hospital and when they come to our appointments… we need to figure out… ‘OK, how we can help you? Do we need to call a porter or do we need to get someone to physically assist you to bring you here?”’ – P12 (Physiotherapist)

### Professional role and identity

This theme describes clinical staffs’ perceptions of their job role and related identity in relation to facilitating appointment attendance, service delivery and patient care.

### Duty of care

Several staff had a strong sense of duty of care in their role as healthcare providers, both in the process of treating patients and facilitating attendance. One physiotherapist described their role as,

> “we’re public servants and we’re here to do a job, and that’s to treat patients.” - P09.

However, the perceptions of the responsibilities constituting duty of care varied, particularly in relation to how to engage with patients. Some staff highlighted that when a patient does not attend an appointment, depending on the pathology (i.e., level of severity of the patient’s clinical condition), there is concern that the patient’s condition may have worsened.

> “…I think as clinicians, we have a duty of care… if you’ve got concerns about a patient and then they’ve got some serious pathology, then you’ll probably keep calling until you get hold of them and say, ‘is everything OK’, because you need to safety net those patients” - P09 (Physiotherapist)

However, there was also an indication that often staff are left wondering if the patient who missed the scheduled appointment is interested in receiving care for their condition.

> “So, when they don’t come, my first concern is, is the patient OK?… When we contact the patients back because they haven’t shown – usually that’s my first question – is the patient OK? Have they had a fall? Has something happened at home? Then obviously the second concern, besides the clinical one would be like ‘OK, is this patient interested in coming and seeing us?’, because if the patient is not interested, it is the cost to the service and basically our time that has been allocated and not used in the best way. And probably the third one will be – is the patient, because he is or is not interested and is not attending, is that going to cause recurrence or any complications?” – P12 (Physiotherapist)

### Mitigation strategies mapped to the TDF

Participants described a range of current and prospective non-attendance mitigation strategies, in addition to related barriers and facilitators associated with their adoption in practice. These were deductively mapped to one of the 14 domains of the TDF with the aim of identifying suggestions for future interventions that could positively impact patient engagement. Seven out of the 14 TDF domains were identified as relevant, including: knowledge; reinforcement; goals; optimism; memory, attention and decision-making; environmental resources and context; and emotions (see Table 2).

**Table 2.**
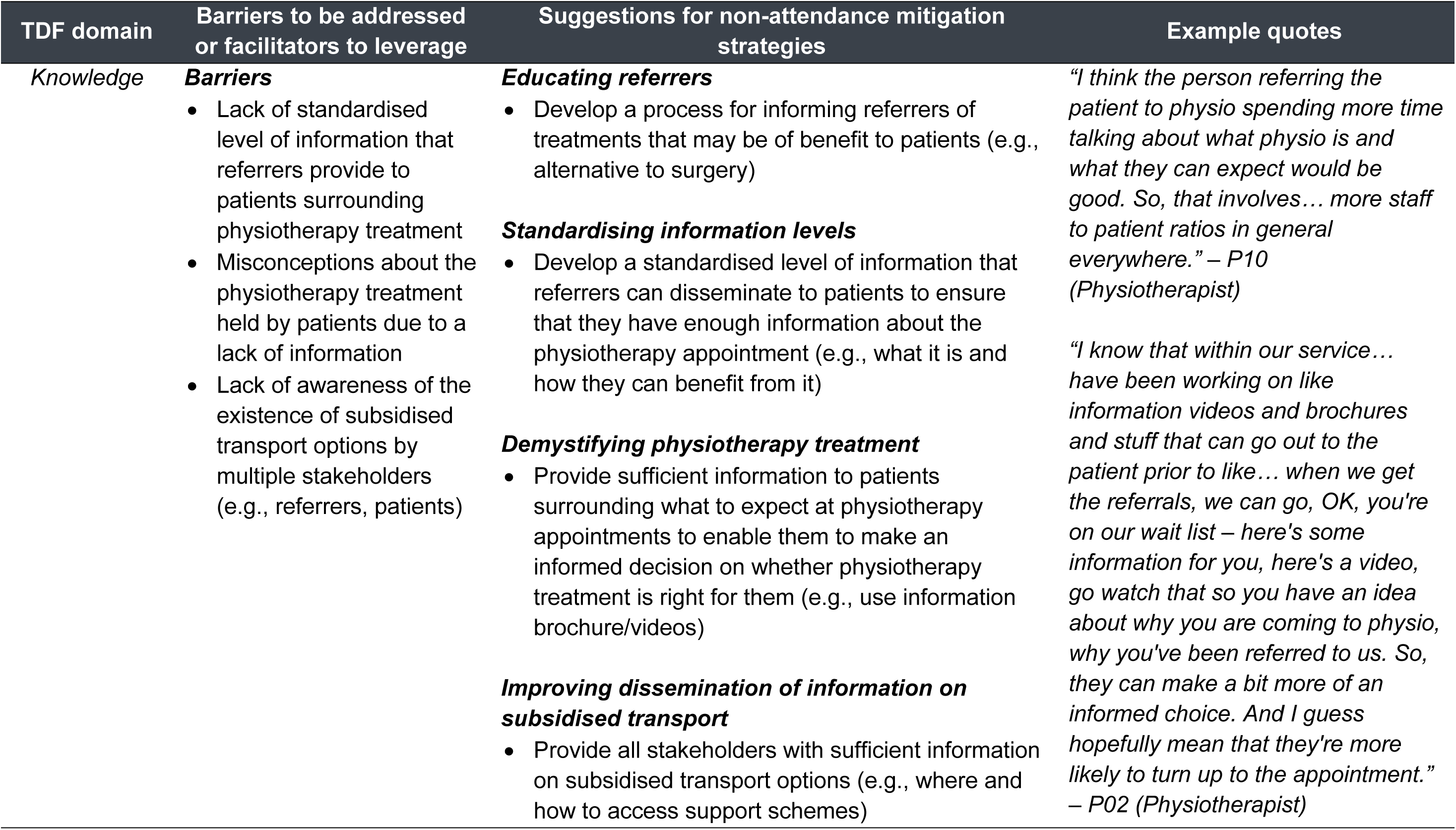
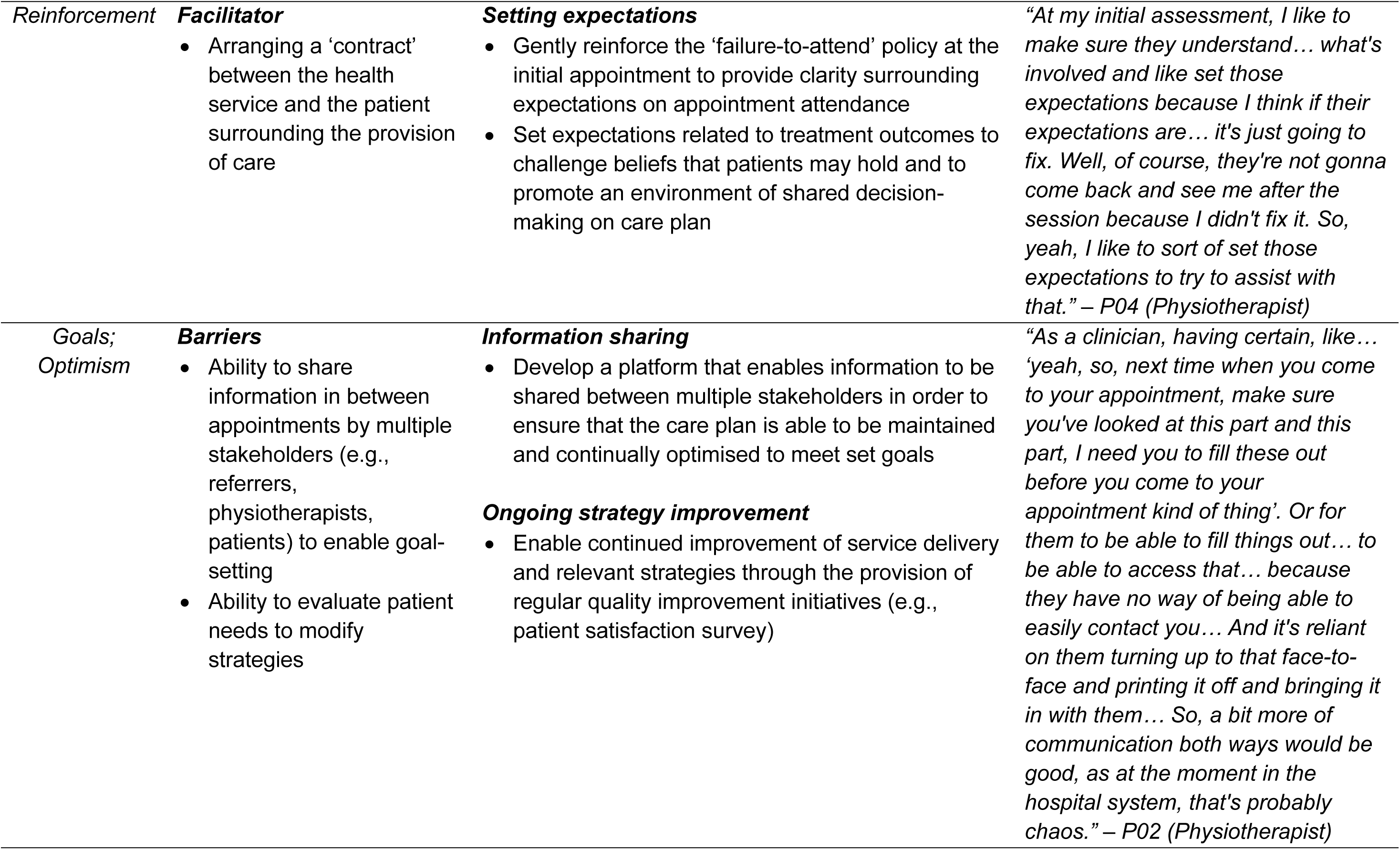
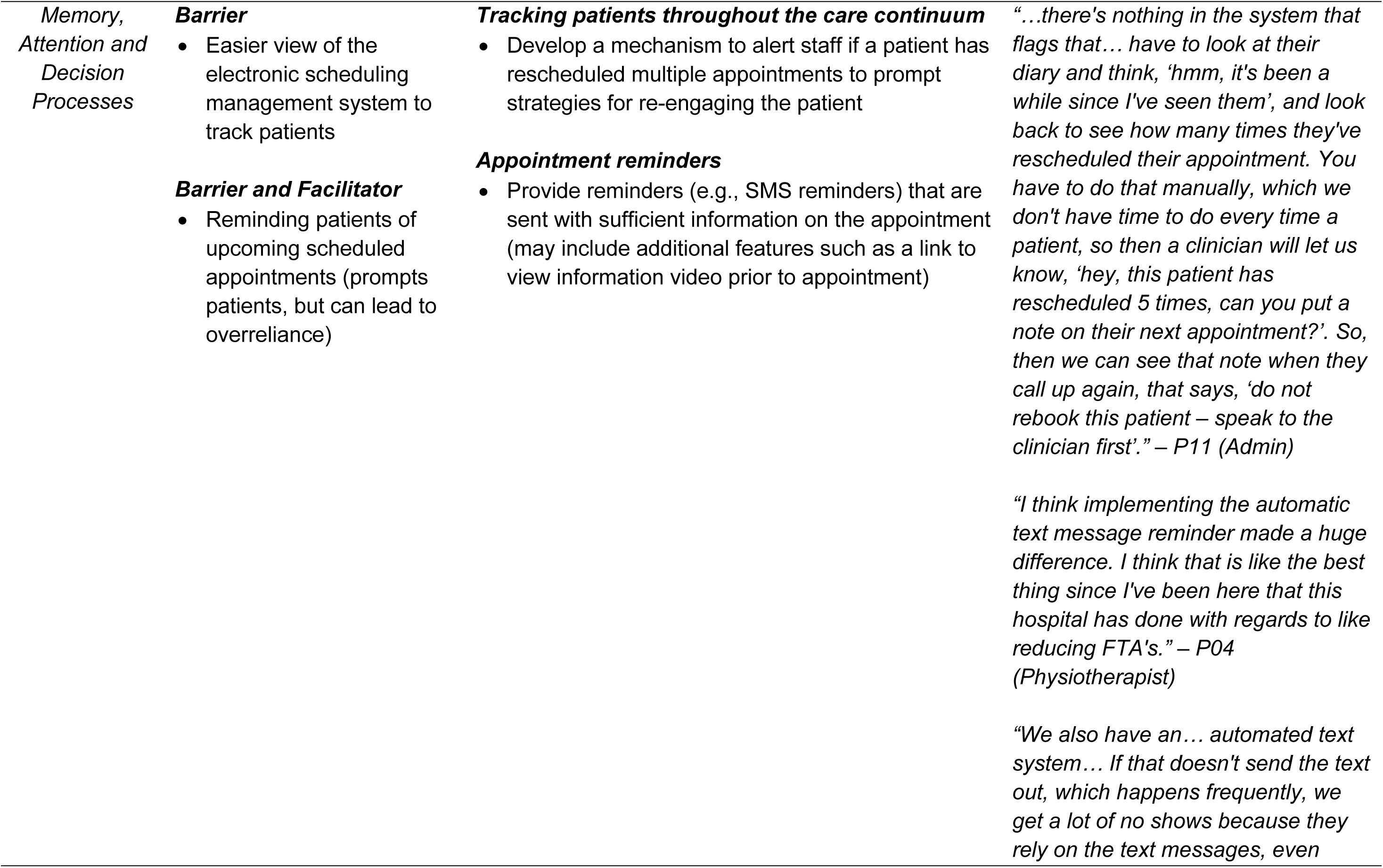
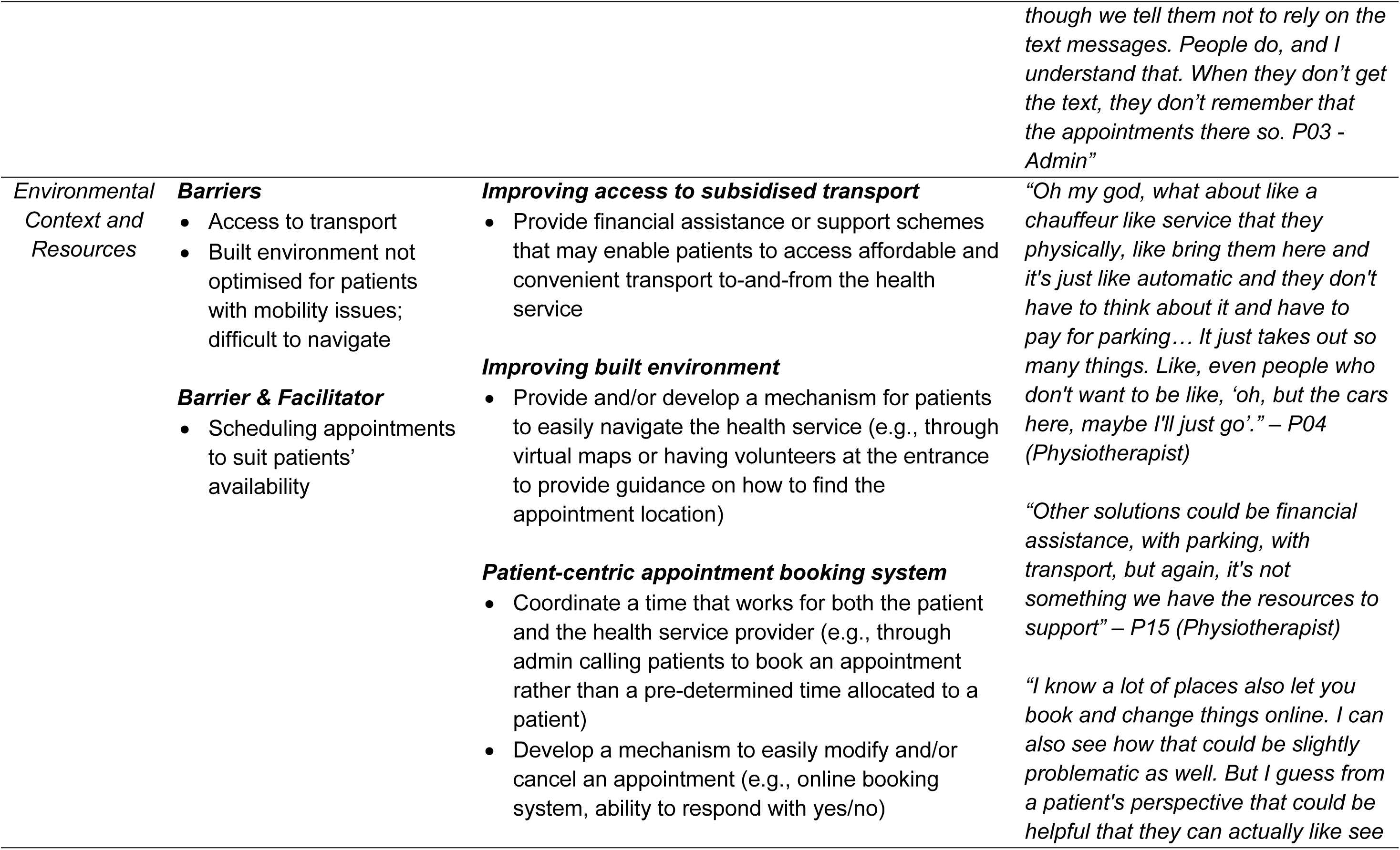
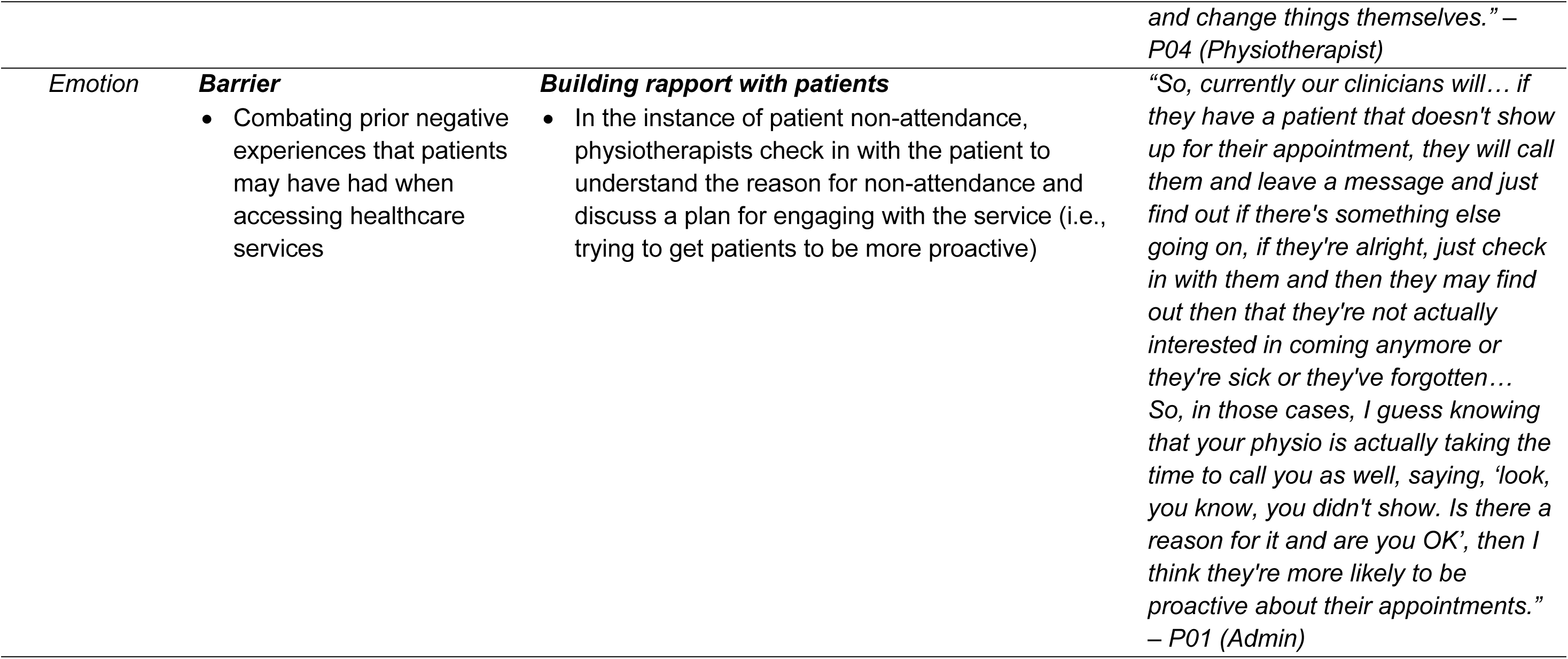
Suggestions for non-attendance mitigation strategies.

## Discussion

Exploring healthcare professional’s views on non-attendance may provide insights into the development and implementation of pragmatic, acceptable and contextually appropriate solutions. Hence, this study sought to explore clinic staff’s thoughts on the impacts of non-attendance, the barriers and facilitators associated with the implementation of non-attendance mitigation strategies, and to identify health service staff-generated solutions to address the perceived barriers and enhance facilitators.

In this study, non-attendance was indicated to have predominantly negative implications for the health service, healthcare provider, and patient. This aligns with findings from other studies that illustrate how non-attendance continues to be an ongoing concern for many healthcare providers. Across the globe, non-attendance puts a strain on healthcare systems’ resources, has the ability to reduce efficiency and quality of service provision, and can disproportionately impact the health of population groups who experience vulnerability.^3,38–40^

Findings from this study suggest an interplay between several different factors that contribute to patient non-attendance. These factors can be considered on a structural or organisational (pertaining to the health system or service), relational (pertaining to the relationship between the provider and patient), or individual (pertaining to the patient) level. Structural factors identified in this study include access to transport, appointment scheduling and flexibility, and built environment. Relational factors include negative experiences, fear of being stigmatised, and level of knowledge and education. Individual factors include perceived benefits and competing commitments (loss of opportunity). These factors often interact with one another and can transcend beyond their designated levels. For example, staff perceived that the types of patients who were more likely to attend an appointment were often those who saw tangible benefits from attending (e.g., those who saw an improvement in their condition). On the flip side, staff expressed that transportation difficulties such as the lack of affordable parking options at the service or lack of transport provision (e.g., availability of someone to drop them off) were potential contributors to non-attendance. Patients experiencing this form of material deprivation may potentially perceive value in attending appointments but be restricted by the lack of access to affordable transport options that would enable them to attend. Some staff indicated that a potential solution to address transportation difficulties could be financial assistance schemes. However, staff emphasised that there appears to be a lack of awareness about these schemes among providers and patients. Thus, exploring ways to better disseminate information on support schemes and whether access to such schemes translates to improved attendance remains a priority for future investigation.

Perception of value may vary depending on the level of knowledge a patient has on the benefits of physiotherapy treatment. Staff indicated that a patient who may not understand why they were referred to physiotherapy may be more likely to not attend an appointment. This is another example of how factors associated with non-attendance may not necessarily exist within one level (e.g., both a relational and individual factor), and thus mitigation strategies will need to consider this complexity. Suggestions for non-attendance mitigation strategies associated with addressing issues surrounding knowledge included educating referrers, standardising information levels that referrers communicate to patients, and demystifying physiotherapy. These suggestions resonate with recommendations in other studies that highlight the need for effort to be placed on improving the mechanisms for communication between healthcare providers and their patients, and between health professions (e.g., referrer and specialists).^4,9,41^ Further, an example of how provider-patient communication may be improved, as identified in this study, includes developing a platform that enables information to be shared between the health service provider and patient in order to ensure that care plans are able to be maintained and continually optimised to meet set goals. This may facilitate better engagement with the health service and give patients the opportunity to take ownership of their condition by providing a means to make informed decisions about the treatment they receive.

The ability to track patients throughout the care continuum was considered by some staff as an integral feature that would enable them to be alerted if a patient had rescheduled multiple appointments. This would provide a means to prompt strategies for re-engaging the patient with the health service rather than having them go unnoticed. This suggested mitigation strategy sheds light on a key concern related to what does and does not constitute non-attendance (i.e., operational definition and classification of non-attendance). Non-attendance has often been characterised as a person who does not attend an appointment, and whose absence may lead to a vacant appointment slot that cannot be filled by other patients.^41,42^ Those who reschedule multiple times or who cancel too late (e.g., 5 minutes before the scheduled appointment time) may also impact the health system in the same way, where a vacant slot is unable to be filled. In addition, both rescheduling and cancelling, unless otherwise specified, may result in inconsistent appointment attendance, which could be equally detrimental to patient health outcomes. Thus, consideration may need to be given to whether the operational definition of non-attendance needs to change, if a subcategory should be created, or if current characterisations should remain. Evaluating current definitions may provide the healthcare system with more precise language to distinguish between different issues being faced by healthcare providers. It may also enable more appropriate metrics to be used for the evaluation of service delivery.

This study had three main limitations. First, the findings from this study would likely be applicable for clinical and administrative staff within a publicly-funded physiotherapy outpatient clinic, or for those working in clinic settings with similar characteristics. Future research may explore other settings (e.g., endocrinology, radiology) and systems (e.g., privately-funded), which may draw other contextually relevant considerations surrounding non-attendance. Second, several findings from this study appear similar to studies that have explored patient’s perspectives on reasons for non-attendance. For instance, patients who found it complicated to reach a healthcare provider either in terms of lack of affordable parking or insufficient transport provision were more likely to fail-to-attend an appointment.^4^ In another study, patients who prioritised appointment attendance were those who attached value to them either because they had healthcare concerns, were symptomatic or recognised the need for regular check-ups.^43^ The same study also found that patients may often be reluctant to attend appointments due to not being aware that they had been referred to a specialist and lacked the understanding of why they may need the referred appointment.^43^ Despite these similarities, it is unknown if the findings would be the same for patients who have attended the clinics involved in this study. Thus, the views of patients on current and prospective non-attendance mitigation strategies remain a key area to explore in future research. Third, it must be noted that this study was a qualitative exploration of healthcare professionals’ perceptions of the many intersecting factors associated with patient non-attendance and potential interventions for addressing this issue. Further empirical work is required to identify causal mechanisms and to test the effectiveness of intervention strategies.

## Conclusion

Non-attendance is a complex phenomenon. For new practices to be implemented and/or existing practices to be altered within healthcare organisations, changes are required collectively between the health system and healthcare providers.^24^ Additionally, to change behaviour, a clear understanding of the barriers and facilitators influencing behaviour in the context in which they occur is required.^24^ This study found that staff acknowledged the complexities of addressing non-attendance and expressed an understanding of the range of circumstances that contribute to patient non-attendance. Staff identified multiple strategies for reducing non-attendance; many of these strategies would require additional resourcing. Research determining the effectiveness, scalability and sustainability of these strategies both in the short- and long-term following adoption into practice remains a priority for future investigation.

## Declarations

### Author contributions

SMS, SN, SM, HC and SK conceived the study. SMS, SN, SM recruited participants and conducted the focus group/interviews. SMS, SN, CM analysed the data. SMS drafted the manuscript. SMS, SN, CM, SM, HC and SK revised and edited the manuscript. All authors read and approved the final manuscript.

### Ethics statement

This study received ethical approval from the Metro South Health Human Research Ethics Committee (HREC/2021/QMS/81605) and written informed consent was obtained from all participants.

## Data Availability

The dataset (which includes individual transcripts) is not publicly available due to confidentiality policies.

## References

1. Molfenter, T. Reducing appointment no-shows: going from theory to practice. Subst. Use Misuse 48, 743–749 (2013).

2. Wolff, D. L. et al. Rate and predictors for non-attendance of patients undergoing hospital outpatient treatment for chronic diseases: a register-based cohort study. BMC Health Serv. Res. 19, 1–11 (2019).

3. Bech, M. The economics of non-attendance and the expected effect of charging a fine on non-attendees. Health Policy 74, 181–191 (2005).

4. Wilson, R. & Winnard, Y. Causes, impacts and possible mitigation of non-attendance of appointments within the National Health Service: a literature review. J. Health Organ. Manag. 36, 892–911 (2022).

5. Campbell, K., Millard, A., McCartney, G. & McCullough, S. Who is least likely to attend? An analysis of outpatient appointment ‘did not attend’(DNA) data in Scotland. Edinb. NHS Health Scotl. (2015).

6. Schwalbe, D., et al. Causes of Patient Nonattendance at Medical Appointments: Protocol for a Mixed Methods Study. JMIR Res. Protoc. 12, e46227 (2023).

7. Lacy, N. L., Paulman, A., Reuter, M. D. & Lovejoy, B. Why we don’t come: patient perceptions on no-shows. Ann. Fam. Med. 2, 541–545 (2004).

8. Ofei-Dodoo, S., Kellerman, R., Hartpence, C., Mills, K. & Manlove, E. Why patients miss scheduled outpatient appointments at urban academic residency clinics: a qualitative evaluation. *Kans*. J. Med. 12, 57 (2019).

9. Akter, S., Doran, F., Avila, C. & Nancarrow, S. A qualitative study of staff perspectives of patient non-attendance in a regional primary healthcare setting. Australas. Med. J. 7, 218 (2014).

10. Husain-Gambles, M., Neal, R. D., Dempsey, O., Lawlor, D. A. & Hodgson, J. Missed appointments in primary care: questionnaire and focus group study of health professionals. Br. J. Gen. Pract. 54, 108–113 (2004).

11. Pyo, J., Lee, W., Choi, E. Y., Jang, S. G. & Ock, M. Qualitative research in healthcare: necessity and characteristics. J. Prev. Med. Pub. Health 56, 12 (2023).

12. Amin, J. et al. Is physiotherapy an underused approach to prevent surgery in selective musculoskeletal disorders? *Bangladesh J*. Med. Sci. 20, 409–413 (2021).

13. Fransen, M. When is physiotherapy appropriate? Best Pract. Res. Clin. Rheumatol. 18, 477–489 (2004).

14. Australian Physiotherapy Association. Value of Physiotherapy in Australia. (2020).

15. Australian Institute of Health and Welfare. Chronic musculoskeletal conditions. Aust. Gov. (2023).

16. Gill, T. K. et al. Global, regional, and national burden of other musculoskeletal disorders, 1990–2020, and projections to 2050: a systematic analysis of the Global Burden of Disease Study 2021. Lancet Rheumatol. 5, e670–e682 (2023).

17. Duckett, S. Getting an initial specialists’ appointment is the hidden waitlist. The Conversation (2018).

18. Tong, A., Sainsbury, P. & Craig, J. Consolidated criteria for reporting qualitative research (COREQ): a 32-item checklist for interviews and focus groups. Int. J. Qual. Health Care 19, 349–357 (2007).

19. Queensland Government. Metro South Health - About Us. (2023).

20. Bearman, M. Eliciting rich data: A practical approach to writing semi-structured interview schedules. Focus Health Prof. Educ. Multi-Prof. J. 20, 1–11 (2019).

21. Noyes, J. et al. Cochrane Qualitative and Implementation Methods Group guidance series—paper 1: introduction. J. Clin. Epidemiol. 97, 35–38 (2018).

22. Thorne, S. Interpretive Description: Qualitative Research for Applied Practice. (Routledge, 2016).

23. Atkins, L. et al. A guide to using the Theoretical Domains Framework of behaviour change to investigate implementation problems. Implement. Sci. 12, 1–18 (2017).

24. Cane, J., O’Connor, D. & Michie, S. Validation of the theoretical domains framework for use in behaviour change and implementation research. Implement. Sci. 7, 1–17 (2012).

25. French, S. D. et al. Developing theory-informed behaviour change interventions to implement evidence into practice: a systematic approach using the Theoretical Domains Framework. Implement. Sci. 7, 1–8 (2012).

26. Avent, M. L., Franks, W., Redmond, A., Allen, M. J. & Naicker, S. Developing an intervention package to optimise the management of vancomycin therapy using theory informed co-design. Res. Soc. Adm. Pharm. (2024).

27. McGowan, L. J., Powell, R. & French, D. P. How can use of the Theoretical Domains Framework be optimized in qualitative research? A rapid systematic review. Br. J. Health Psychol. 25, 677–694 (2020).

28. Thorne, S. & Darbyshire, P. Land mines in the field: A modest proposal for improving the craft of qualitative health research. Qual. Health Res. 15, 1105–1113 (2005).

29. Thompson, A. P., MacDonald, S. E., Wine, E. & Scott, S. D. Understanding parents’ experiences when caring for a child with functional constipation: interpretive description study. JMIR Pediatr. Parent. 4, e24851 (2021).

30. Birt, L., Scott, S., Cavers, D., Campbell, C. & Walter, F. Member checking: a tool to enhance trustworthiness or merely a nod to validation? Qual. Health Res. 26, 1802–1811 (2016).

31. Motulsky, S. L. Is member checking the gold standard of quality in qualitative research? Qual. Psychol. 8, 389 (2021).

32. Duff, J., Bowen, L. & Gumuskaya, O. What does surgical conscience mean to perioperative nurses: An interpretive description. Collegian 29, 147–153 (2022).

33. Fereday, J. & Muir-Cochrane, E. Demonstrating rigor using thematic analysis: A hybrid approach of inductive and deductive coding and theme development. Int. J. Qual. Methods 5, 80–92 (2006).

34. Lumivero. NVivo (Release 1.6.1). (2017).

35. Thompson Burdine, J., Thorne, S. & Sandhu, G. Interpretive description: A flexible qualitative methodology for medical education research. Med. Educ. 55, 336–343 (2021).

36. King, N., Brooks, J. & Horrocks, C. Interviews in qualitative research. (2018).

37. Sellars, M., White, B. P., Yates, P. & Willmott, L. Medical practitioners’ views and experiences of being involved in assisted dying in Victoria, Australia: A qualitative interview study among participating doctors. Soc. Sci. Med. 292, 114568 (2022).

38. Bedford, L. K., Weintraub, C. & Dow, A. W. Into the storm: a mixed methods evaluation of reasons for non-attendance of appointments in the free clinic setting. *SN Compr*. Clin. Med. 2, 2271–2277 (2020).

39. George, A. & Rubin, G. Non-attendance in general practice: a systematic review and its implications for access to primary health care. Fam. Pract. 20, 178–184 (2003).

40. Alturbag, M. Factors and Reasons Associated With Appointment Non-attendance in Hospitals: A Narrative Review. Cureus 16, (2024).

41. Mohammed Selim, S., Kularatna, S., Carter, H., Bohorquez, N. G. & McPhail, S. M. Digital Health Solutions for Reducing the Impact of Non-Attendance: A Scoping Review. Health Policy Technol. 100759 (2023).

42. Mbada, C. E. et al. Impact of missed appointments for out-patient physiotherapy on cost, efficiency, and patients’ recovery. Hong Kong Physiother. J. 31, 30–35 (2013).

43. Copeland, S., Muir, J. & Turner, A. Understanding Indigenous patient attendance: a qualitative study. Aust. J. Rural Health 25, 268–274 (2017).

